# How to improve access to TB care for the nomads? Review of barriers and enablers for Sahel nomadic populations

**DOI:** 10.1101/2022.05.06.22274767

**Authors:** Hugues Asken Traore, Roxane Chaverondier, Adjima Combary, Adama Diallo, Salifou Ouedraogo, El hadj Kane, Mohamedou Koita, Assao Mourtala, Sina Adamou, Marie Sarr, Nafissatou Toure, Tacko Aly Ba, Oumar Abdelhadi, Abderramane Abdelrahim, Bakary Konaté, Yaya Ballayira, Fatima Camara, Madelon Roij, Corinne Simone Merle

## Abstract

Tuberculosis (TB) control in nomadic populations represents a major public health problem in sub–Saharan Africa.

Barriers and enablers of TB care for nomads were identified through a literature review and survey conducted among the National TB Programs (NTPs) of six Sahelian countries: Burkina Faso, Chad, Niger, Mali, Mauritania, and Senegal. A conceptual framework was developed. Data retrieved from twenty-eight peer-reviewed papers or collected through the survey were regrouped in 5 categories: health system related factors, socioeconomic factors, cultural, political and environmental factors.

The large distance between nomadic camps and health care facilities and the absence of TB-specific programmatic interventions for nomads were the main barriers identified. The establishment of a multi-ministerial national committee in charge of nomadic populations, the mapping of nomadic transhumance roads, the identification of gaps in health service provision and community engagement for defining fit for purpose solutions are key elements to improve TB control in nomadic population.

Some countries in the region successfully implemented interventions to overcome the barriers to TB care. These interventions should be more widely shared to inform other countries for the development of appropriate strategies for which community engagement is essential.

## Introduction

Tuberculosis (TB) in nomadic populations forms a major public health problem. Undiagnosed and untreated patients face a heavy burden of morbidity and mortality and they are also an important reservoir and continue to transmit TB (1). The term “nomad” here refers to nomadic and semi-nomadic communities, including pastoral and agro-pastoral communities. It does not refer to migrants and populations displaced for economic, religious, or political reasons or in the context of armed conflict.

The world population of nomads and semi-nomads is estimated at 50-100 million, of whom 60% live in Africa (1). Between 20 and 30 million of nomadic pastoralists live in the Sahel region. Obtaining precise demographic country-level data from nomadic communities is challenging, given their seasonal cross-border mobility (2). Among nomadic pastoralists we note a high TB prevalence due to living conditions, including promiscuity, and lack of TB control measures. TB remains underdiagnosed. Without treatment, transmission remains active. TB control measures are not in place as access to care is poor (1, 3).

Apart from barriers experienced by the general population (lack of infrastructure and trained staff, inadequacies of the health system, particularly with regards to free TB care, stigmatization of TB patients, and treatment-related side effects) (4), nomads experience additional sociocultural barriers to TB care. No previous study reviewed these barriers and described interventions that could overcome them.

The purpose of this study was to review published evidence and get National Tuberculosis Programmes (NTPs) perception on (i) the barriers encountered by nomads to use TB services and (ii) on interventions piloted to overcome these barriers in the Sahel region in Africa.

## Methods

### Study design

We reviewed the literature on barriers to TB service utilization and measures or interventions that were used to overcome these barriers among nomadic populations in the Sahel region. We also conducted a survey to gather the perception and experiences of NTP representatives of six Sahel countries.

### Literature search strategy and eligibility criteria

The search strategy involved: (1) electronic database searching (MedLine, Elsevier), (2) a web search (Google Scholar), (3) searching websites from United Nations system such as WHO, FAO, UN refugees Agency; non-governmental organizations such as Club du Sahel et de l’Afrique de l’Ouest and the World Bank website, and (4) snowballing by screening the reference lists of retrieved articles. The publication date was not an exclusion criterium. We only selected publications in English and French. We used search strings that combined keywords. **Table 1** shows how the string was constructed for the search on Medline, using Pubmed. It was adapted to fit searches on other websites.

**Table 1:** Keys words for building search strings used during the literature review.

|  |  |
| --- | --- |
| # 1 | nomad OR nomadic OR “semi-nomadic” OR pastoralists OR agropastorolism OR pastoralism |
| # 2 | Sahel OR “Burkina Faso” OR Chad OR Mali OR Mauritania OR Niger OR Senegal OR Africa [MeSH] OR "Africa, Central"[Mesh] OR "Africa, Western"[Mesh] |
| # 3 | Health OR “health care” OR “health access” OR “health care delivery” OR “tuberculosis”<br>OR<br>MeSH terms: Tuberculosis/drug therapy ; |
| # 4 | 1 AND 2 AND 3 |

### Data collection

All articles identified based on the four search strategies were imported into the reference management software, Zotero (version 5.0.18) for screening and managing records, as well as to remove duplicate references. If screening did not exclude records based on title or abstract, the full text was assessed.

### Data extraction and summary

The following data was extracted from selected records: (1) barriers to the utilization of TB services, (2) measures or interventions used to overcome barriers, and (3) recommendations for future measures or interventions, and listed in separate tables, stratified by type of barrier (health system barriers; socio economic, cultural or political barriers; environmental barriers)

### Data collection process

First the principal investigators (HT and RC) did an electronic search of Medline, Elsevier, and Google scholar. Duplicates were excluded. After reading the title and abstract, the full text of articles addressing the topic of interest was assessed. Eligibility for inclusion in the review (include the disagreements) were decided or treated by CM based on reading the full text.

### Survey among NTPs of 6 Sahel countries

This survey was conducted within the framework of the West African Regional Network for Tuberculosis (WARN-TB) and Central African Regional Network for Tuberculosis (CARN-TB) networks. Access to TB care for the nomadic population was discussed during a regional meeting and the survey was proposed to all NTPs. self-administered questionnaire was sent by e-mail to the NTP teams in Burkina Faso, Mali, Mauritania, Niger, Senegal and Chad who expressed interest to contribute. They were also contacted by phone and/or email for explaining the questionnaire content and facilitating questionnaire completion. Some NTP representatives were contacted a second time when further clarification was needed.

A conceptual framework was developed. Data retrieved from twenty-eight peer-reviewed papers or collected through the survey were regrouped in 5 categories: health system related factors, socioeconomic factors, cultural, political and environmental factors **(Figure 1)**.

**Figure 1:**
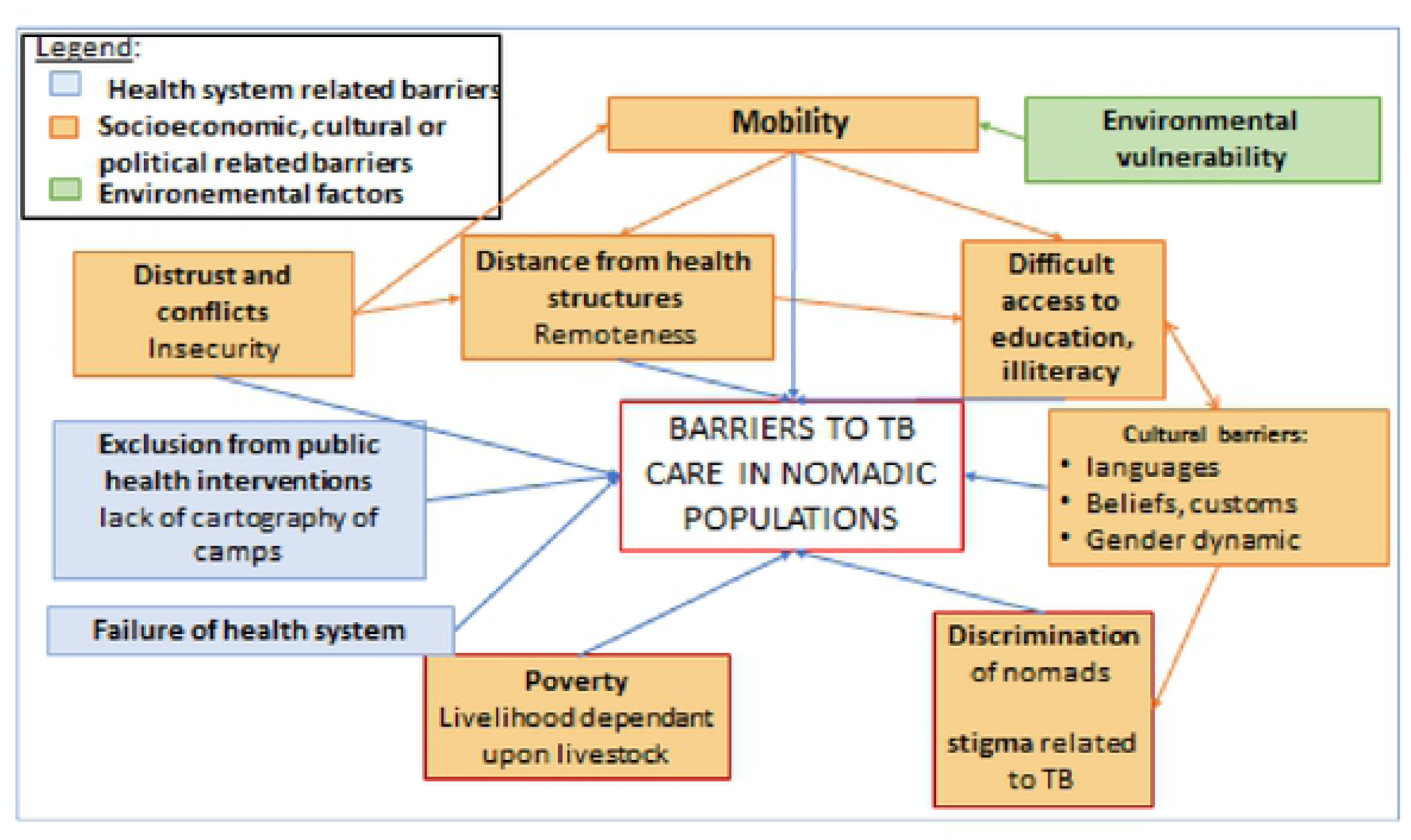
Conceptual Framework.

## Results

Through the electronic search of Medline, Elsevier, and Google scholar we identified 314 articles, of which 308 unique articles (figure 1). After reading the title and abstract, the full text of 118 articles was assessed for eligibility. Finally, 28 studies were included in the review (**figure 2**). The studies were conducted in the following countries (numbers of peer-reviewed papers): South Africa (1), Ethiopia (7), Kenya (3), Mali (2), Niger (2) Nigeria (7), Uganda (1), Somalia (1), Sudan (1), Chad (3). Additional information was collected through the contribution of the NTPs of Burkina Faso, Mali, Mauritania, Niger, Senegal, and Chad.

**Figure 2:**
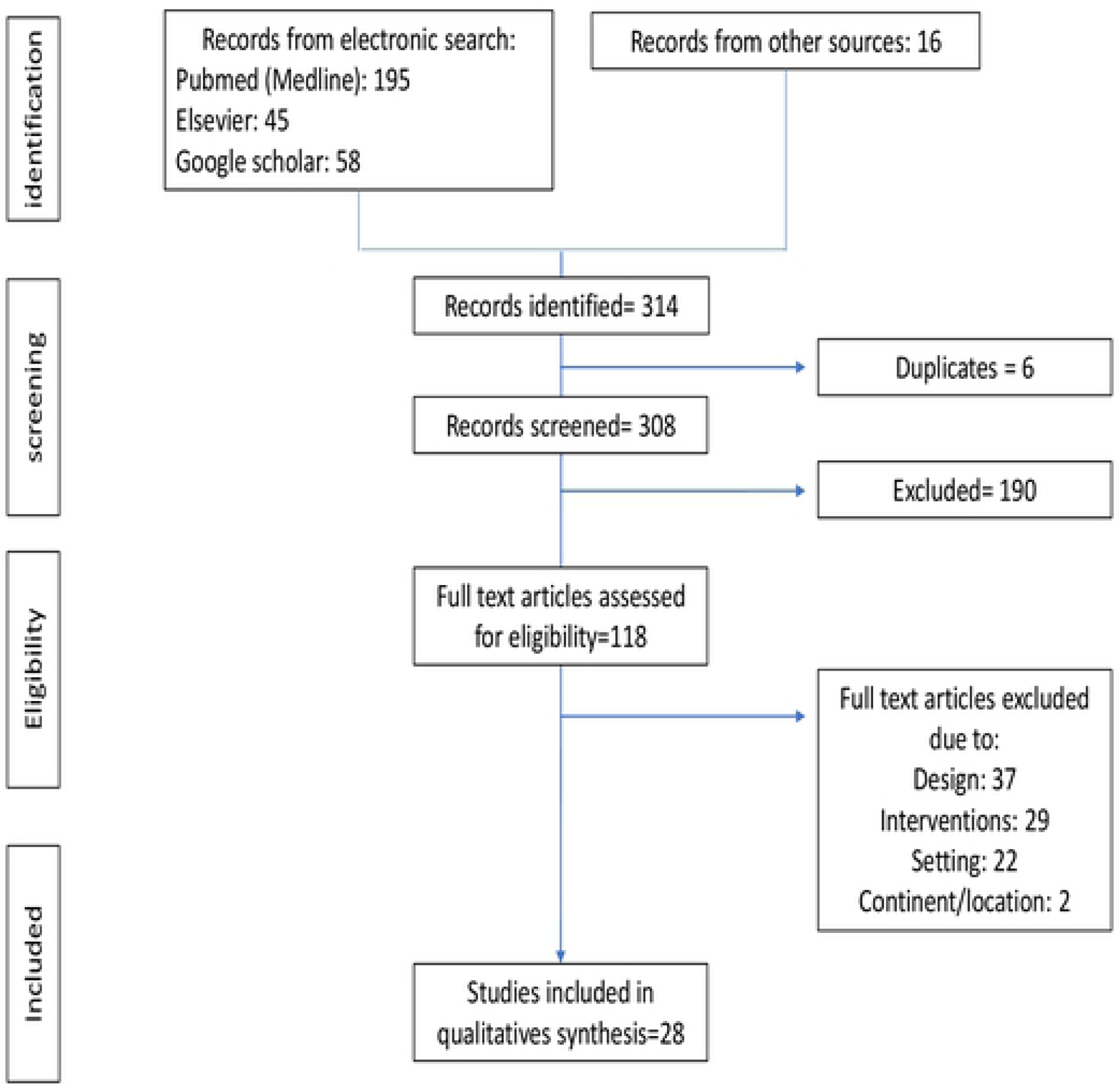
flowchart showing the process of articles selection.

Barriers to TB diagnosis and treatment of TB in the nomadic populations evoked by the NTPs of the different countries were numerous, belonged to different domains and were similar to those mentioned in the literature **(table 2)**. Health system barriers included lack of health infrastructure, lack of trained personnel, and dysfunctions of the health system. Another structural barrier was a geographical one, and related to the distance between communities and health facilities. Some barriers were specific for nomadic populations, and were related to their culture, religion, beliefs, customs, level of education, and mobility.

**Table 2:** Barriers to TB services in nomadic populations or reported by NTPs of Sahel countries or shown in the literature.

| Barriers |  | Countries/NTP |  |  |  |  |  | Literature review |
| --- | --- | --- | --- | --- | --- | --- | --- | --- |
|  |  | Burkina Faso | Mali | Niger | Mauritania | Senegal | Chad |  |
| Health system barriers | Lack of demographic and health data | x |  |  |  | x |  | Laokri *et al.* 2013 (38) |
|  | Lack of trust between nomads and health services |  |  |  | x |  |  | Rass *et al.* 2018 (22),<br>Brieger *et al.*, 1997 (47)<br>Okeibunor *et al.* 2013 (68) |
|  | Lack of communication between health workers and nomads |  | x |  | x |  |  | Dao *et al.* 1994 (67)<br>Brieger *et al.* 1998(68) |
|  | Lack of community health care workers on transhumance roads and corridors |  |  |  | x |  |  |  |
|  | Lack of mobile health teams |  |  |  | x |  |  | Gele *et al.* 2009 (63)<br>Storla *et al.* 2008 (66)<br>Imperato *et al.* 1975 (43) |
|  | Limited access to screening and treatment services |  | X | x | x | x | x | Laokri *et al.* 2013 (38) |
|  | Lack of early TB detection strategy in nomadic populations (delayed TB diagnosis results in advanced TB disease, which is more difficult to manage at primary health care level) |  | x | x | x | x | x | Sheik-Mohamed *et al.* 1999 (11)<br>Gele *et al.* 2010 (69)<br>Okeibunor *et al.* 2007 (70) |
|  | Lack of treatment follow-up strategy for nomadic tuberculosis patients with the consequence of non-compliance with treatment follow up appointments, including side effects of TB treatment (with a large number of patients being lost to follow-up) |  | x |  |  | x |  | Kisambu *et al.* 2014 (85)<br>Gele *et al.* 2009 (63)<br>Storla *et al.* 2008 (66) |
|  | Underdetection in districts with nomadic populations making treatment monitoring<br>difficult |  |  |  |  | x |  | Gele *et al.* 2009 (63) |
|  | Stigma among<br>health worker |  |  |  | x |  |  | Okeibunor *et al.* 2013<br>(47)<br>Storla *et al.* 2008 (66)<br>Sima *et al.* 2017 (72)<br>Sima *et al.* 2019 (77)<br>Zegeye *et al.* 2019(78) |
|  | Lack of a<br>centralized<br>institute<br>responsible for the<br>management of<br>specific needs of<br>nomadic<br>populations |  |  |  |  |  |  | Lô *et al.* 2016 (84) |
|  | Lack of experience<br>carrying out TB-specific<br>interventions<br>among nomads | x | x |  | x | x |  |  |
| **Socio economic,**<br>**cultural or**<br>**political barriers** | Insecurity | x | x | x |  |  | x |  |
|  | Mobility / no fixed<br>place of residence | x | x | x | x | x | x | Leclerc *et al.* 2011(65) |
|  | Way of life<br>(languages,<br>beliefs, customs,<br>gender dynamic) |  |  | x |  | x |  | Bechir *et al.* 2004 (73) |
|  | Poverty | x | x | x | x | x | x | Mugisha *et al.* 2002<br>(80) |
|  |  |  |  |  |  |  |  | Blench *et al.* 2001 (81)<br>Domenech *et al.* 2013(82) |
|  | Poor access to knowledge and education | x | x | x | x | x | x | Gele *et al.* 2009 (63)<br>Storla *et al.* 2008 (66)<br>Dao *et al.* 1994 (67)<br>Brouwer *et al.* 1998 (71) |
| **Environnemental barriers** | Distance | x | x | x | x | x | x | Dao *et al.* 1994 (67)<br>Brieger *et al.* 1998(68) |
|  | Lack of precise maps of the location of the transhumance camps | x | x | x | x | x | x | John *et al.* 2015 (21)<br>Brieger *et al.* 1997 (68)<br>Gidado *et al.* 2014 (90) |
|  | Desert territory |  | x | x | x |  | x |  |

The summary of the measures and interventions to overcome barriers, as reported by the NTPs of the target countries and/or the literature is shown in **table 3**. A large number of nomads carry out seasonal transhumance, and their mobility makes the provision of medico-social services complex. Most interventions essentially aim to overcome problems linked to seasonal transhumance and the mobility of nomadic populations.

**Table 3:**
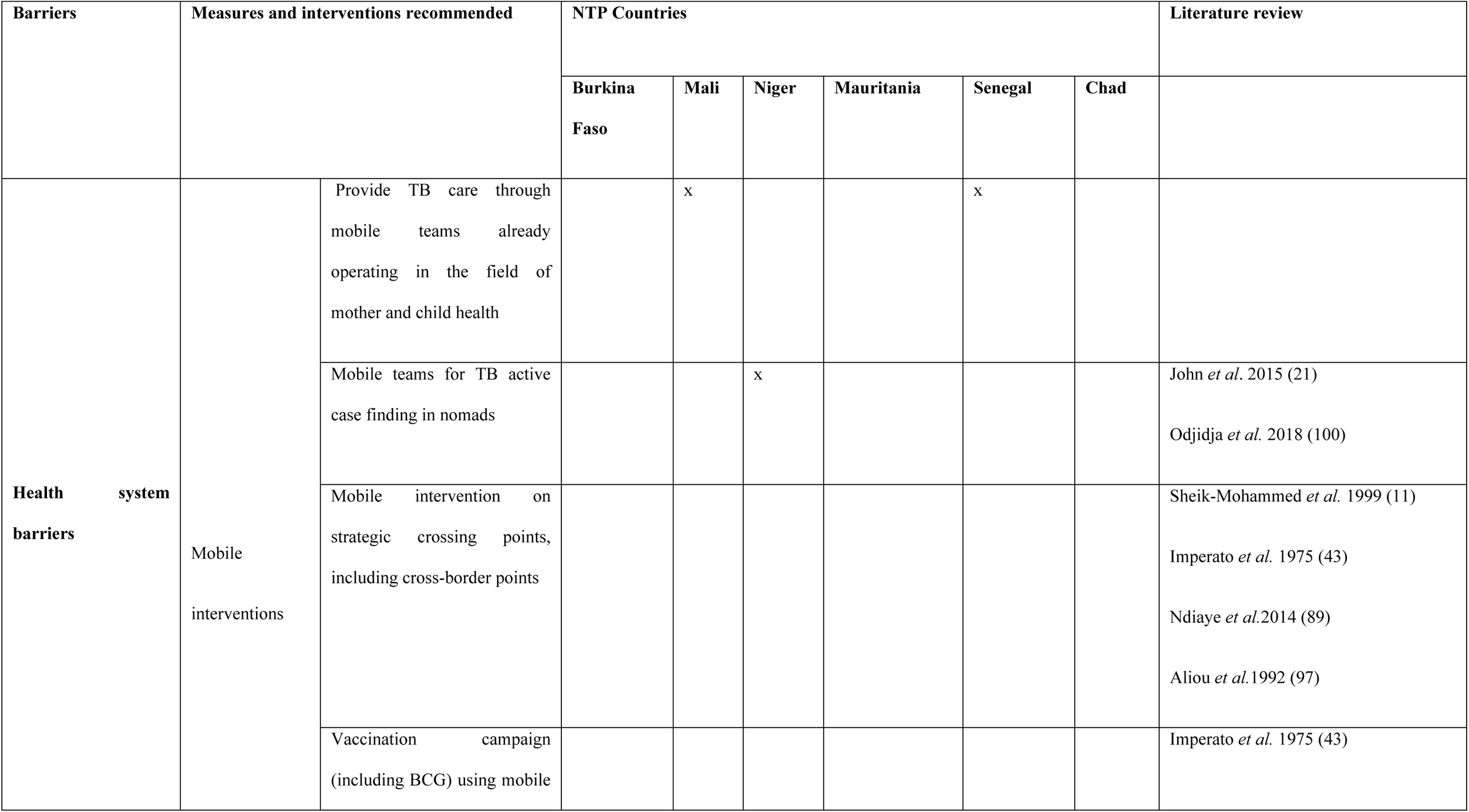
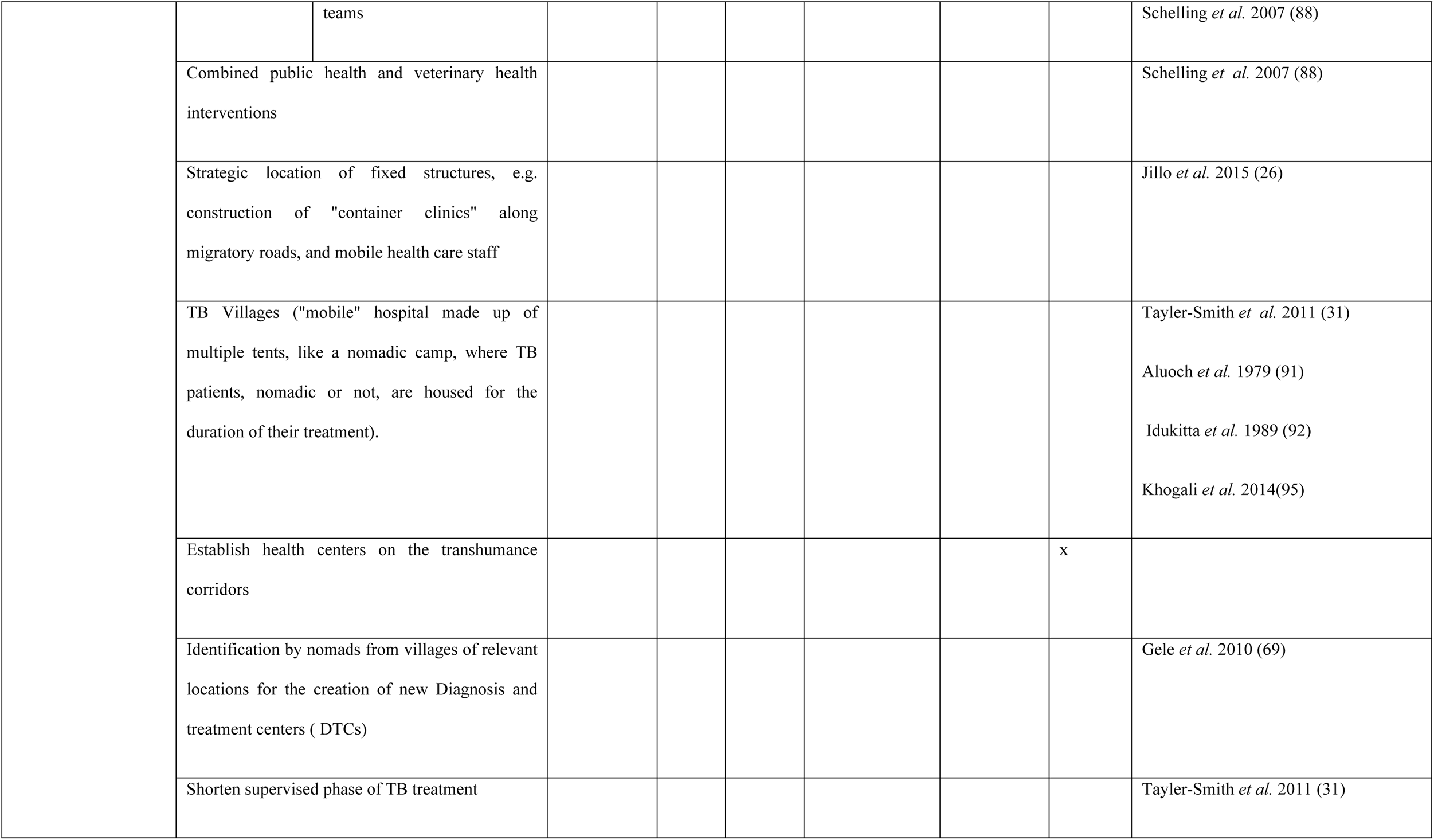
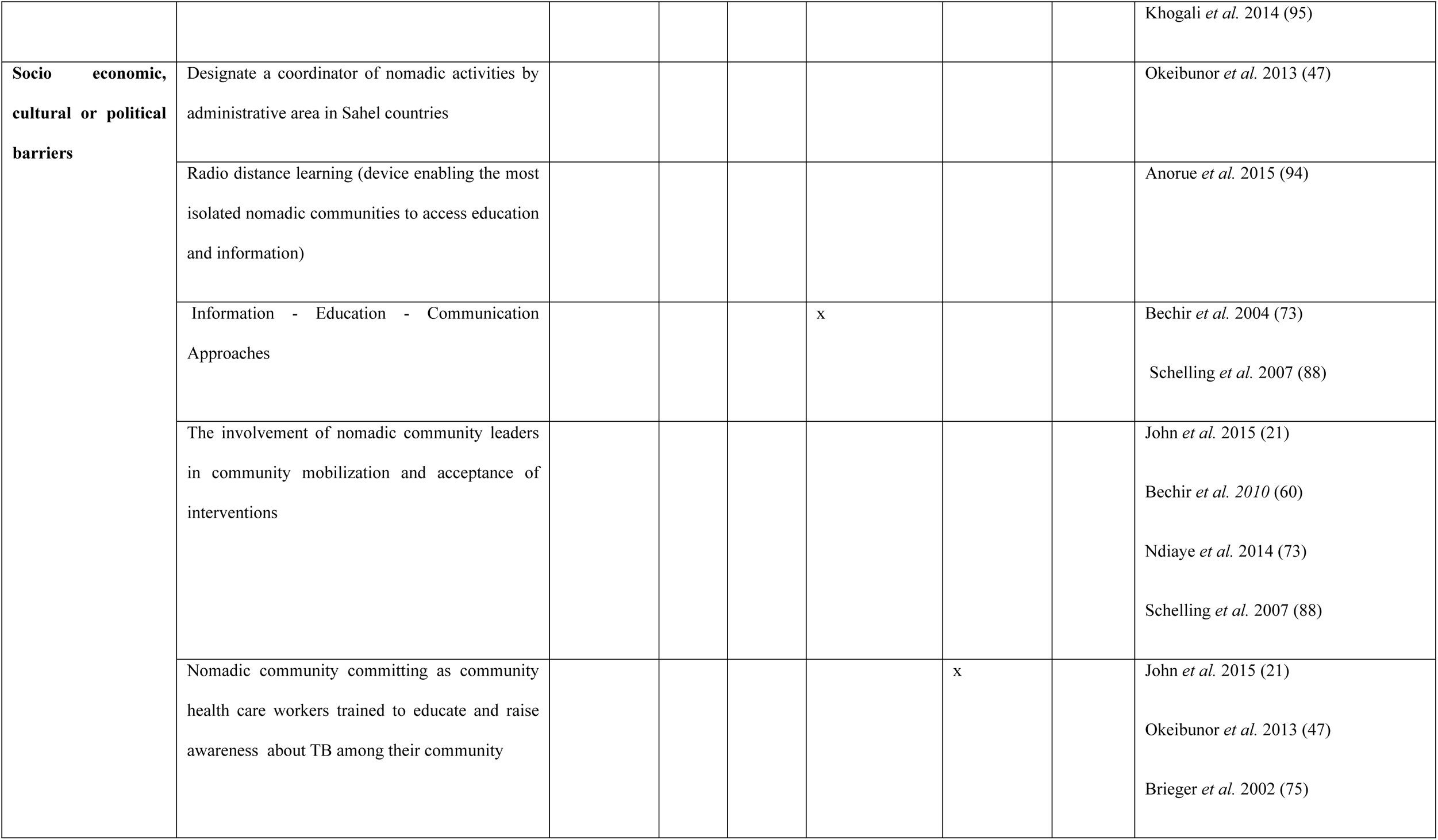
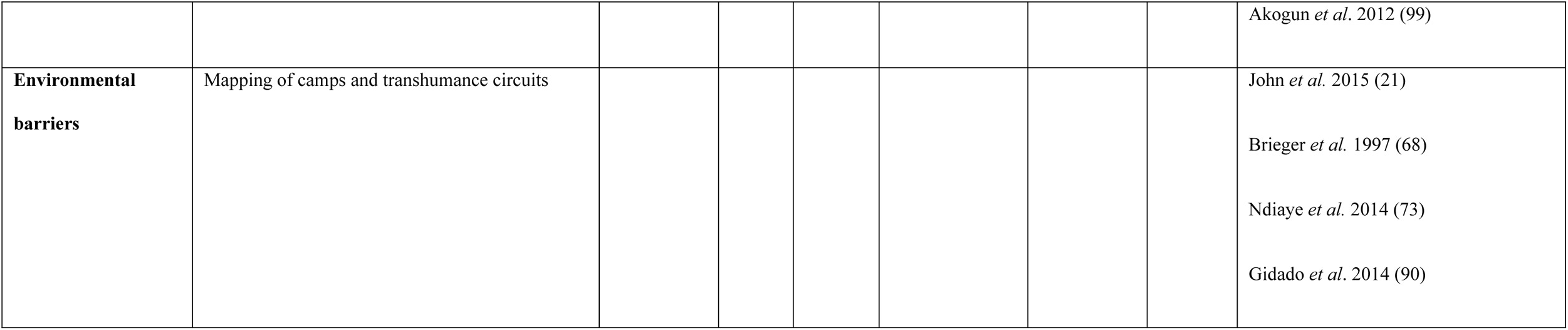
Measures and interventions to overcome barriers to TB services in nomadic populations or reported by NTPs of Sahel countries or shown in the literature.

The strategic axes for interventions or measures recommended by the NTPs and also reported in the literature **(table 4)** cover both health system barriers, geographical and nomadic population specific barriers. Recommendations include the roll out of mobile interventions, such as mobile clinics, innovative approaches such as the establishment of “TB Village”, and above all the involvement of target communities through training and education.

**Table 4:**
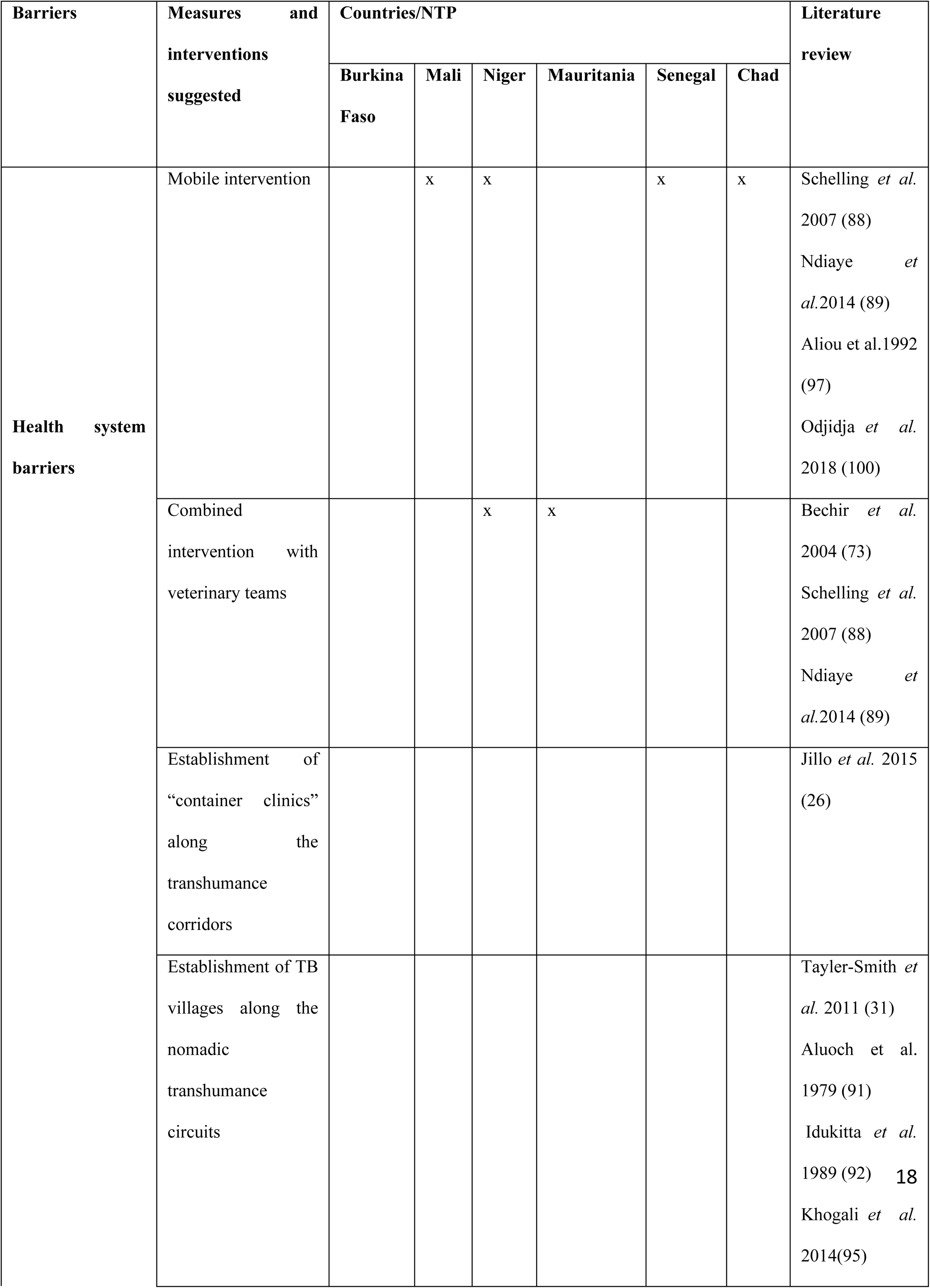
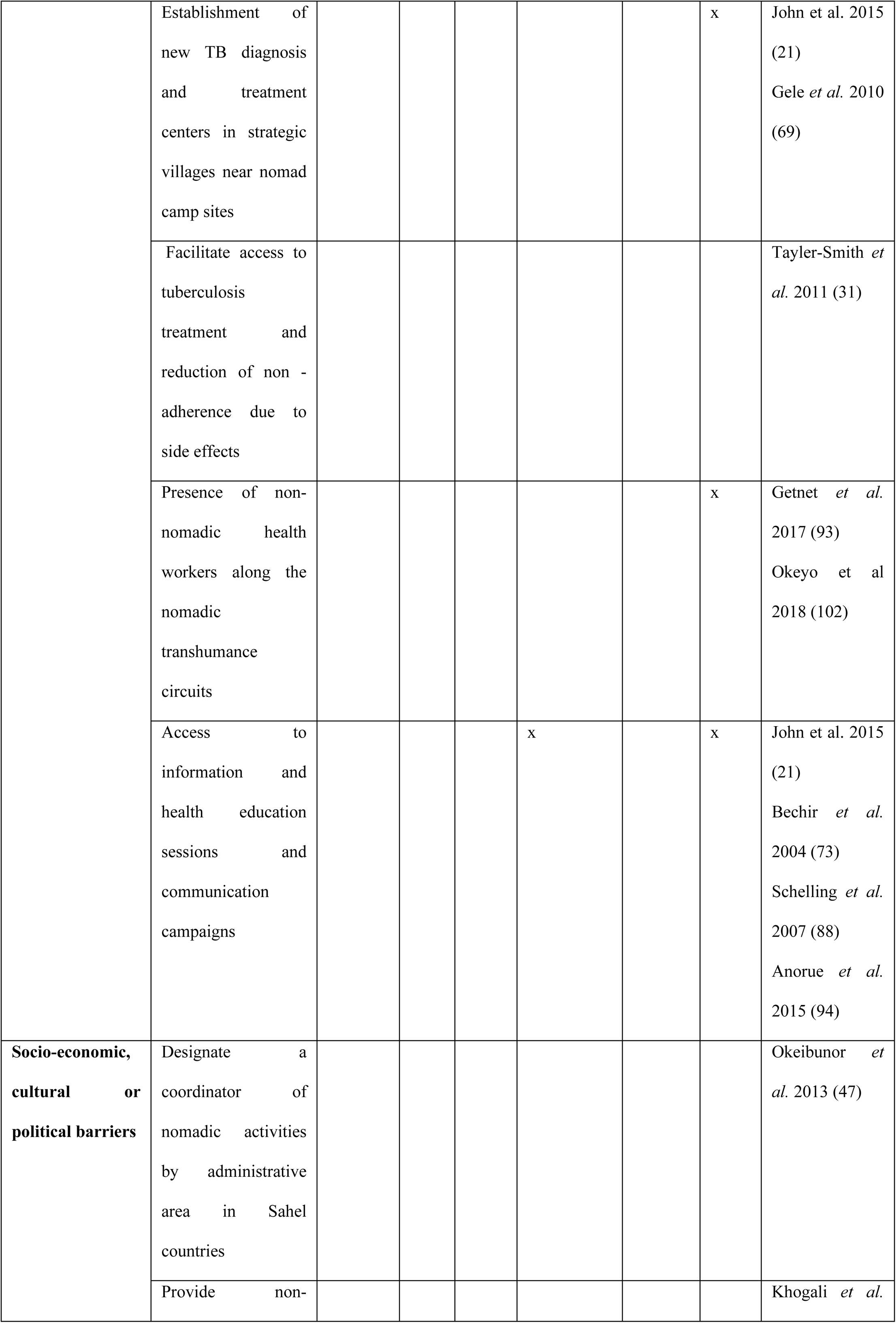
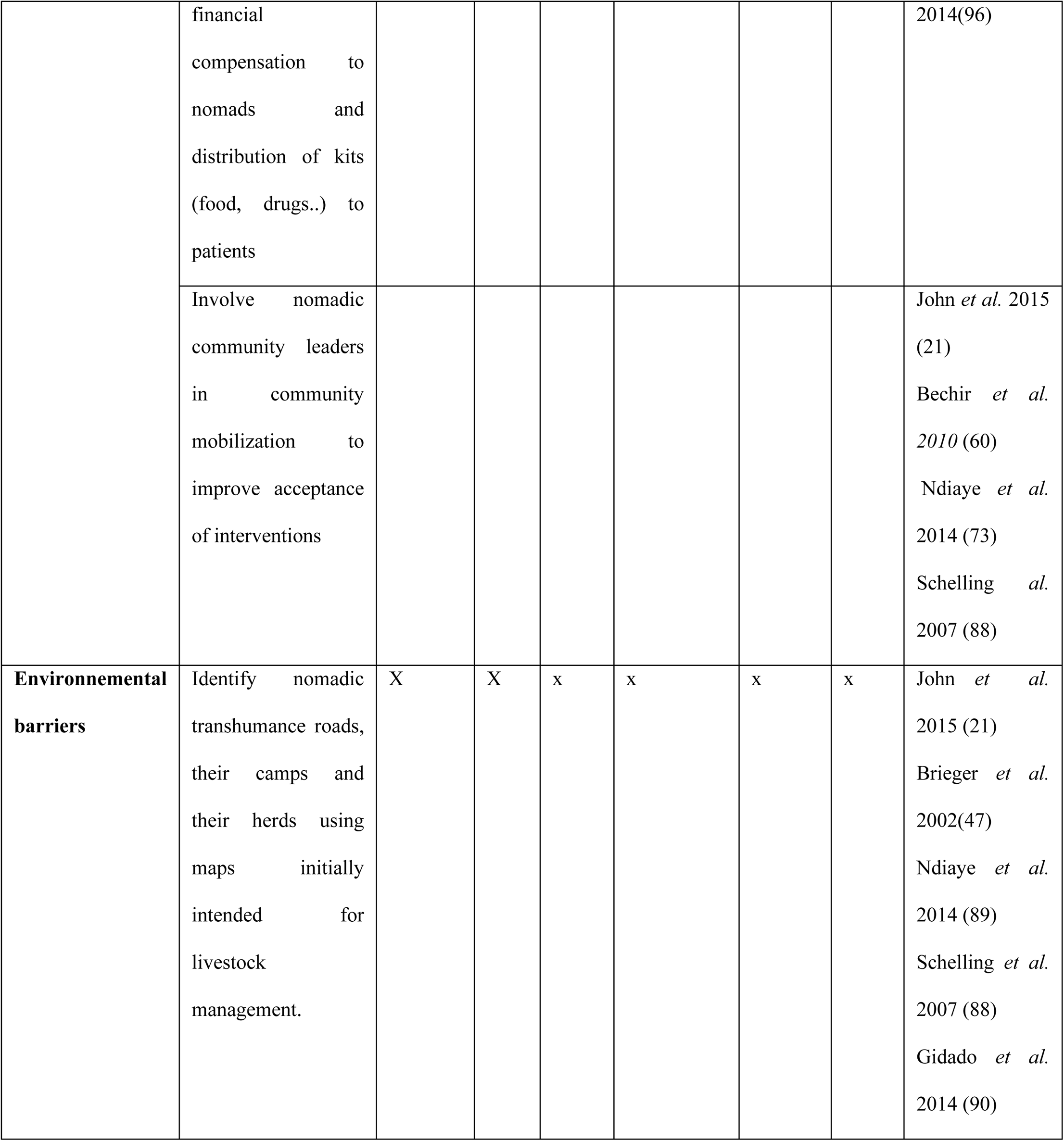
Recommendations for measures and interventions to overcome barriers to TB services in nomadic populations or reported by NTPs of Sahel countries or shown in the literature.

## Discussion

This review analyzed the barriers and interventions for improving access of nomadic populations to TB health care. Based on the review of 28 scientific articles and on the responses received from NTPs of six (06) Sahelian countries in West and Central Africa, we got an overview of barriers to TB care among the nomadic population. This review also helped us to better understand what is needed to be improved and what was piloted already in the region. So far, there is no other study which summarized the barriers to TB care and interventions conducted to improve access of nomads to TB care in Sahelian countries.

Our study shows that nomads face many barriers to accessing TB care. One of the main barriers is the large distance between nomad camps and health care facilities, making access to health care difficult (67, 68). Moreover the nomadic population is often not reached by TB-specific programmatic interventions (11, 63, 66, 69, 70, 85) and is often stigmatized by health care providers (47, 66, 72, 77, 78). In addition to these main barriers, the mobile lifestyle of nomads, the highly prevalent illiteracy and the insecurity regarding desert areas place of transhumance forms a barrier for effective health care (65, 73, 80, 81).

Frequently implemented interventions for nomadic population relied on mobile interventions (11,21, 43, 85, 97, 100) such as TB active case findings in nomads by mobile teams, sometimes combined with another health or veterinary interventions. Schelling et al. (2007) (43) also conducted a vaccination campaign (including BCG) using mobile teams in Chad, which resulted in complete vaccination of 10% of targeted nomadic children, compared to zero percent previously. Similarly, Imperato et al (1969)(72) showed than during immunization campaigns against smallpox in Burkina Faso, Mali and Mauritania that the cattle market was an important gathering point that allowed the vaccination of many nomads, merchants and traders, usually always on the move and rarely found in the villages during national vaccination campaigns. This strategy seems very interesting for carrying out awareness-raising activities and TB diagnostic activities but does not provide any ideas for improving BCG vaccination of nomadic infants, who are traditionally absent from the markets. Similarly, the problem of monitoring TB treatment would not be solved by this strategy as it stands. In addition, questions of costs could arise; indeed Sheik-Mohammed et al (39) in Botswana showed that for the same level of care, the operating costs of mobile teams would have been 8 times higher than those of dispensaries. Conversely, the intervention reported by Odjidja et al. (2018)(51) report the use of mobile clinics as the most cost-effective for meeting the goals of their infectious disease control program among pregnant Sudanese pastoralists. Finally, Bechir et al. (2004)(102), Ndiaye et al. (2014)(53) and Schelling et al. (2007)(43) establish that intersectoral collaboration between public health and veterinary health services can reduce the costs of mobile interventions carried out with nomads by sharing, when possible, human and material resources. Using this strategy in Chad, Schelling et al. (2007)(43) were able to reduce the logistical costs of certain rounds by up to 15% during their vaccination campaign.

In order to bring care closer to nomads, “container clinics” were set up along transhumance roads. This was described by Zinsstag J and al (2004) (26) at Chad and by Jillo and al. (2015)(70) at Kenya. The clinic opens when nomadic pastoralists come nearby. A statistically significant difference was observed before and after the intervention for the number of medically assisted deliveries, going from 5.6% to 17.7% in the population studied. A better knowledge of the importance of antenatal visits was also observed. On the other hand, this project has generated little change in practices on the number of prenatal consultations carried out by nomadic women. This strategy, according to Zinsstag J and al (2004) (26), applied at tuberculosis, would be interesting to implement because it responds to the problems of mobility and distance from health care structures, and could allow an improvement in the diagnosis and follow-up of TB in nomads.

TB Villages which are “mobile” hospitals made up of many tents, similar to nomadic camps, were also proposed to accommodate TB patients (both nomadic as non-nomadic) for the duration of their treatment (92, 95). This approach was also carried out in Kenya by Aluoch (1979) (113) and by Idukitta et al. (1989) (73). The TB villages were named “Manyatta” which means mobile house. These ‘TB villages’ had good treatment success rates, 80% for the Ethiopian“TB village” of Tayler-Smith et al (2011)(34), and 81% treatment success for the evaluable Kenyan ‘Manyatta’ of Idukitta et al. It should be noted that the above-mentioned approaches require the patient to take the personal step of going to the “TB village”. It therefore falls under “passive screening” for TB. It implies prior effective communication campaigns to make known the existence of the structure to potential patients and to encourage them to come to the TB village, which was done effectively in the program of Tayler-Smith et al. (2011)(34).

Regarding the costs of implementing such projects, the “TB village” of Tayler-Smith et al. (2011) (34) benefited from the support of the NGO MSF and their human and logistical resources. This collaboration between MSF and the NTP of Chad seems to have greatly contributed to the quality and good results of this project in terms of success rate of TB treatment. The overall cost of caring for a TB patient and their caregiver, including accommodation, food and materials, during the entire TB treatment period was estimated to be around 315 US$ and 390 US$ per patient treated for the 6-month and 8-month respectively.

In the “TB village” or “Manyatta” of Aluoch (1979) (113), food was not provided and the camp was built by the patients themselves, which meant that the costs of such a project were not prohibitive to its implementation at this time

Interventions with nomadic communities’ engagement were also carried out such as raising awareness of TB by education (21, 47, 75, 99) and involvement of nomadic community leaders in TB education and community mobilization (21, 60, 73, 88). Community directed interventions allow decentralization of care to reach the most isolated rural areas. Thus, reducing the distance between the source of information and the beneficiaries improves access to knowledge about the pathology in question [Brieger et al. (2002)(96), Okeibunor et al. (2013)(49), Akogun et al. (2012)(52). John et al. (2015)(69), trained volunteers within the nomadic community to continue active TB screening activities over the long term. The involvement of nomadic community leaders is central to community mobilization and acceptance of interventions, as observed in Nigeria by John et al. (2015)(69) and in Chad by Ndiaye et al. (2014)(53), Schelling et al. (2007)(43) and Bechir et al. (2004)(102). Community-led interventions and CHW training are measures that also address many cultural barriers (Brieger et al. (2002)(96), Okeibunor et al. (2013)(49), Akogun et al. (2012)(52).

A large number of measures and solutions undertaken in the countries of the Sahel or in other African regions to respond to the many specificities of the management of TB in nomadic populations have been identified. Each project carried out with these populations tries to best achieve its objectives according to the local socio-economic context and the means that the level of development of the country allows it to deploy.

Several other interventions are suggested in the literature or by NTPs and could be implemented in Africa. These are primarily focused on identifying nomadic transhumance roads and camps (for example using maps initially intended for livestock management) (21, 47, 88, 89, 90) and integration of TB interventions in already existing “advanced and mobile care strategies” in the field of mother and child health and vaccination campaigns in nomadic areas (88, 89, 97, 100). These approaches have been documented as efficient strategies in similar socio-economic contexts (53, 43,70) and it is necessary to verify their feasibility in the nomadic populations of the Sahel countries. Indeed, certain obstacles to carrying out these interventions can hardly be overcome without being accompanied by a certain level of development in the countries of interest, such as the presence of paved roads in remote regions, or even peace and security in the Sahel region. Finally, it is expected that engaging various governmental and non-governmental partners would make the best use of the resources and field data already available in the target countries and would make it possible to gain logistical and economic efficiency. Like The Global fund fight against AIDS, Tuberculosis and Malaria (GF) which has already engaged on this path in Niger and Chad. Funding has been granted to the country for the evaluation of differentiated strategies for improving nomads’ access to tuberculosis care.

In summary, the non-sedentary lifestyle of nomads and the absence of a health policy focused on this special group are the cause of numerous barriers to access to TB care for nomads. With the growing interest of donors (as the World Health Organisation) in improving TB screening and treatment in hard-to-reach populations such as nomads, many NTPs will have to reflect and / or implement interventions on this subject.

### Implication

To effectively respond to the needs in terms access to healthcare for vulnerable populations in these regions, governments of the Sahelian countries need to invest in training of qualified personnel and equipment of healthcare infrastructures. An intersectoral partnership on the ministry level (for example a work group between Ministry of Health, Ministry of Education, Ministry of Livestock, Ministry of Environment) has to be created and a better transboundary cooperation has to be established. Moreover, the establishment of a network of field partners (NGO, pastoral organization, associations) remains necessary to have a greater impact of identified interventions.

### Strengths and limitations

The main strength of the current study is that it shows an extensive overview of relevant literature. To provide a more complete overview of barriers and interventions for nomadic access to health care in Africa, this review was triangulated with data obtained from the NTPs. Documents from different programs were retrieved supporting the validation of findings.

Most studies did not compare one intervention with another. It is not yet clear which intervention would work best. Moreover, data transmitted by NTPs were reported by NTP managers, based on their experience. As data were not collected from patients, we cannot conclude which barriers are being experienced by nomads themselves.

### Futures research

Future research among the nomads is important, in particular qualitative studies. Here are the key elements that need to be focused on: (1) The needs of different nomads groups in terms of education, information and access to health care to related to TB, (2) The priorities for the nomads in terms of health structures cartography in each countries: Health centers in strategic locations, the transhumance routes and link those two maps, (3) The potential of a the involvement of the community for TB activities and (4) The relevance of the use of mobile teams or fixed structures regarding to a given nomadic groups in order to have available health structures compatible with mobility patterns. Public health actions targeting the eradication of TB are part of a more general health and social problem and and it is even a greater challenge f its actions are to meet the priorities and essential needs of nomads.

## Conclusion

This study gave us an overview of the obstacles to TB care for the nomadic population and provided a better understanding of what would be needed to do to overcome them. The following approach could be envisaged by countries:

1. To establish an intersectoral partnership with representatives of the Ministry of Health, Ministry of Education, Ministry of Livestock, Ministry of Environment, etc. with transboundary cooperation to discuss specific health and non-health issues of the nomads
2. To create a national network of field partners implementing activities for the benefit of the nomads with that local and international NGOs, pastoral organization to better mutualize resources
3. To document geographical gaps for TB care with mapping TB care offer in terms of availability of TB centre on the transhumance road followed by the nomads
4. To engage the nomadic community in a problem-solving demarche with (i) discussing the gaps found with the mapping, (ii) informing them of potential solutions experimented by other and (iii) for defining which solutions could be adapted to their context. The following aspects should be discussed

- What is the current knowledge on TB of the community, and which are the needs in terms of education and information related to TB?
- Would the community be interested in being involved in TB screening and/or care activities?
- Should there be specific mobile teams for addressing TB care needs or should the already existing structures be strengthened to better address nomads needs regarding TB care?
- Could e-Health be an interesting tool to facilitate TB care activities?
5. To pilot the strategy and document through the conduct of implementation research the effectiveness, feasibility, acceptability and eventually cost of the piloted strategy in order to inform the scale up of the proposed strategy

## Supporting information

Annex S1_Questionnaire for NTP

## Data Availability

All data related the findings are already provided of the submitted article.

## Acknowledgments

Our thanks go to Dr TOM Decroo from the Institute of Tropical Medicine Antwerp (Belgium) for his support and guidance in the process of writing this article. Thanks to Dr Isidore TRAORE from the University of Ouagadougou (Burkina Faso) for his review of this article

